# COVID-19 vaccine effectiveness among healthcare workers in Portugal: results from a hospital-based cohort study, December 2020 to November 2021

**DOI:** 10.1101/2022.01.07.22268889

**Authors:** Vânia Gaio, Adriana Silva, Palmira Amaral, João Faro Viana, Pedro Pinto Leite, Carlos Matias Dias, Irina Kislaya, Baltazar Nunes, Ausenda Machado

## Abstract

**Introduction:** Healthcare workers (HCW) were amongst the first prioritized for COVID-19 vaccination but data on COVID-19 vaccine effectiveness among HCW is still limited. This study aims to estimate the COVID-19 vaccine effectiveness (VE) against SARS-CoV-2 symptomatic infection among HCW from Portuguese hospitals.

**Methods:** In this prospective cohort study, we analysed data from HCW (all professional categories) from two central hospitals in the Lisbon and Tagus Valley and Centre regions of mainland Portugal between December 2020 and November 2021. VE against symptomatic SARS-CoV-2 infection was estimated as one minus the confounder adjusted hazard ratios by Cox models considering age group, sex, presence of chronic disease and occupational exposure to patients diagnosed with COVID-19 as adjustment variables.

**Results:** During the 11 months of follow up, the 2213 HCW contributed a total of 1950 person-years at risk and 171 SARS-CoV-2 events occurred. The COVID-19 incidence rate for unvaccinated HCW was 348.7 per 1000 person-years while for fully vaccinated HCW was 43.0 per 1000 person-years. We observed a VE against symptomatic SARS-CoV-2 infection of 73.9% (95% CI: 26.2–90.8%) for complete vaccination status.

**Conclusion:** This cohort study found a high COVID-19 VE against symptomatic SARS-CoV-2 infection in Portuguese HCW, which is in concordance with previous studies from other countries. Monitoring of VE in this HCW cohort continues during the winter 2021/2022 to evaluate potential VE decay and booster vaccine effect.

## Introduction

The Coronavirus disease 2019 (COVID-19) pandemic is an ongoing health issue of great concern since December 2019. As of November 11^th^, 2021, 82.20.993 million cases and over 1.496.017 deaths had been reported to the WHO European region (1). After implementation of unprecedented non-pharmacological public health measures, including confinement orders in many countries, COVID-19 vaccines were developed and were made available. Since then, vaccination has proved to be an essential tool to reduce transmission of the severe acute respiratory syndrome coronavirus 2 (SARS-CoV-2), as well as severe illness and COVID-19 related mortality (2).

Health Care Workers (HCW) are essential to ensure healthcare for patients in a pandemic context (3, 4). In Portugal, as in many other countries, healthcare workers were amongst the first prioritized for COVID-19 vaccination as they are at increased risk for exposure to SARS-CoV-2 due to their close contact with COVID-19 patients and, additionally, HCW can transmit the infection to susceptible patients at high risk of severe COVID-19. According to the Portuguese vaccination plan, vaccination of HCW started on December 27, 2020, mainly with mRNA vaccine (Comirnaty) and AdV vaccine Vaxzervria (AstraZeneca) (5).

Despite the high efficacy of these vaccines, many factors can affect their performance in a real-world situation, outside the strict setting of clinical trials, which supports the importance of observational studies to assess vaccines effectiveness (VE) (6). Most observational studies initially described high VE for available vaccines. However, VE decreases over time both due to the emergence of new variants, namely the Delta variant, but also due to waning of protection (7–9).

Results from cohort studies conducted in the USA, Denmark and Italy estimate VE among HCW above 80% (10–14). However, these studies reported short follow-up periods, and longer follow-up studies among HCW are still limited, especially regarding the period of the Delta variant predominance. This study aims to estimate the COVID-19 vaccine effectiveness among hospital HCW against symptomatic disease between December 2020 and November 2021, in Portugal.

## Methods

### Study design and population

We developed a cohort study (with retrospectively collected information regarding vaccination and previous infection) that targeted hospital HCW (all professional categories), eligible for vaccination against COVID-19, without contraindications for vaccination, who consented to participate and were employed by 2 hospitals in the Lisbon and Tagus Valley and Centre regions of Portugal (CHTV - Centro Hospitalar Tondela-Viseu and CHLO - Centro Hospitalar Lisboa Ocidental). Only HCW workers with available complete information regarding their vaccination status (1^st^ and 2^nd^ dose data) were included.

### Procedures and collected information

All procedures implemented in this cohort study were based on the Guidance Document “Cohort study to measure COVID-19 vaccine effectiveness among health workers in the WHO European Region” (15). All HCW were invited to participate by an email sent by the occupational health service of each hospital. After accepting to participate and signing a written consent, each HCW answered an enrolment questionnaire implemented in the REDcap platform (16). Recruitment information included sociodemographic, health status, vaccination history, previous SARS-CoV-2 infection, occupational and community exposure and preventive behaviours. Self-reported individual COVID-19 vaccination status was also confirmed by the occupational health service of each hospital. Further, on a weekly basis, participants answered a follow-up questionnaire (through the REDcap platform) which included questions on symptoms and COVID-19 testing in the previous 7 days. The active follow-up started in May 2021, but information regarding RT-PCR (reverse transcription polymerase chain reaction) testing was retrospectively obtained to all participants by the occupational health service of each hospital (from 27 December 2020, the date on which vaccination started, until the end of the study period on 30 November 2021). RT-PCR testing was performed in hospitals whenever a HCW reported any symptom compatible with COVID-19, in each week.

### Exposure and Outcome definitions

An individual was considered vaccinated 14 days after full vaccination (receiving all doses recommended in the product characteristics). An individual was considered unvaccinated if he or she had not received any dose of COVID-19 vaccine. An individual was considered partially vaccinated 14 days after receiving the first dose and until receiving the second dose.

The outcome of the study was laboratory confirmation by RT-PCR of symptomatic SARS-CoV-2 infection according to the World Health Organization suspected case definition (17). Considering that the vaccination of health care professionals had already started, this information was collected retrospectively, whenever available in the registries of the occupational health services.

### Statistical analysis

Participants characteristics at baseline were described according to the vaccination status (unvaccinated, partially vaccinated and fully vaccinated). We estimated the COVID-19 symptomatic infection rates per 1,000 person-years for each level of vaccination exposure. VE was computed as one minus the confounder-adjusted hazard ratio for symptomatic infection, estimated by time-dependent Cox regression (18) with time-dependent vaccine exposure, adjusted for confounding using 7-day periods as strata, as previously published (19). Additionally, age group (18-35 years/ 36-50 years/ 51-70 years), sex (male/female), self-reported chronic disease (Yes/No) and occupational exposure to COVID-19 diagnosed patients (Yes/No) were also considered as confounding factors. Statistical analysis was performed in R version 4.0.5 (R Foundation, Vienna, Austria).

### Ethical considerations

Written informed consent was obtained from all study participants, after being informed that participation was voluntary and that he/she can withdraw from the study without justification at any time and without consequences, particularly with regard to the offer of vaccination. All data were anonymised before statistical analysis by the occupational health services of each hospital. This study protocol was approved by the ethics committee of the Instituto Nacional de Saúde Doutor Ricardo Jorge (date of approval. 16/04/2021), and by the ethics committees of the two participating hospitals, namely by the ethics committee of CHTV (Ref.06/16/04/2021, date of approval 16/04/2021) and CHLO (Ref. 2141, date of approval 19/05/2021).

## Results

### Participant’s characterization

Among the 2213 participants, 81% (n=1777) were females, 45% (n=973) were aged between 36 and 50 years old, 30% (n=417) declared to have at least one chronic disease and 17% (n=231) reported working directly with COVID-19 patients (**Table 1**). Most participants were fully vaccinated (n=2112, 95%) and only a few remained partially vaccinated (n=90, 4%) or unvaccinated (n=11, 0.5%) at the end of the study period.

**Table 1.** General characteristics of the health care workers, according to their vaccination status.

| Characteristics | Total<br>(n=2213) | Unvaccinated<br>(n=11) | Partially vaccinated<br>(n=90) | Vaccinated<br>(n=2112) |
| --- | --- | --- | --- | --- |
| **Age group (n=2143)** | n (%) | n (%) | n (%) | n (%) |
| 18-35 | 532 (24.8) | 1 (12.5) | 24 (26.7) | 507 (24.8) |
| 36-50 | 973 (45.4) | 5 (62.5) | 45 (50.0) | 923 (45.1) |
| 51-70 | 638 (29.8) | 2 (25.0) | 21 (23.3) | 615 (30.1) |
| **Sex (n=2206)** |  |  |  |  |
| Female | 1777 (80.6) | 9 (81.8) | 75 (83.3) | 1693 (80.4) |
| Male | 429 (19.4) | 2 (18.2) | 15 (16.7) | 412 (19.6) |
| **Chronic Disease**<br>**(n=1405)** |  |  |  |  |
| Yes | 417 (29.7) | - | 23 (28.0) | 394 (29.8) |
| No | 988 (70.3) | 3 (100) | 59 (72.0) | 926 (70.2) |
| **Occupational exposure to**<br>**diagnosed COVID-19**<br>**patients (n=1356)** |  |  |  |  |
| Yes | 231 (17.0) | - | 12 (15.0) | 219 (17.2) |
| No | 1125 (83.0) | 3 (100) | 68 (85.0) | 1054 (82.8) |

### Evolution of vaccination status and events over the study period

The majority of the HCW were fully vaccinated during the first trimester of 2021 (**Figure 1**). Mostly were vaccinated with the Comirnaty (n=1750) and Vaxzevria (n=449) vaccines (Spikevax vaccine uptake was residual).

**Figure 1.**
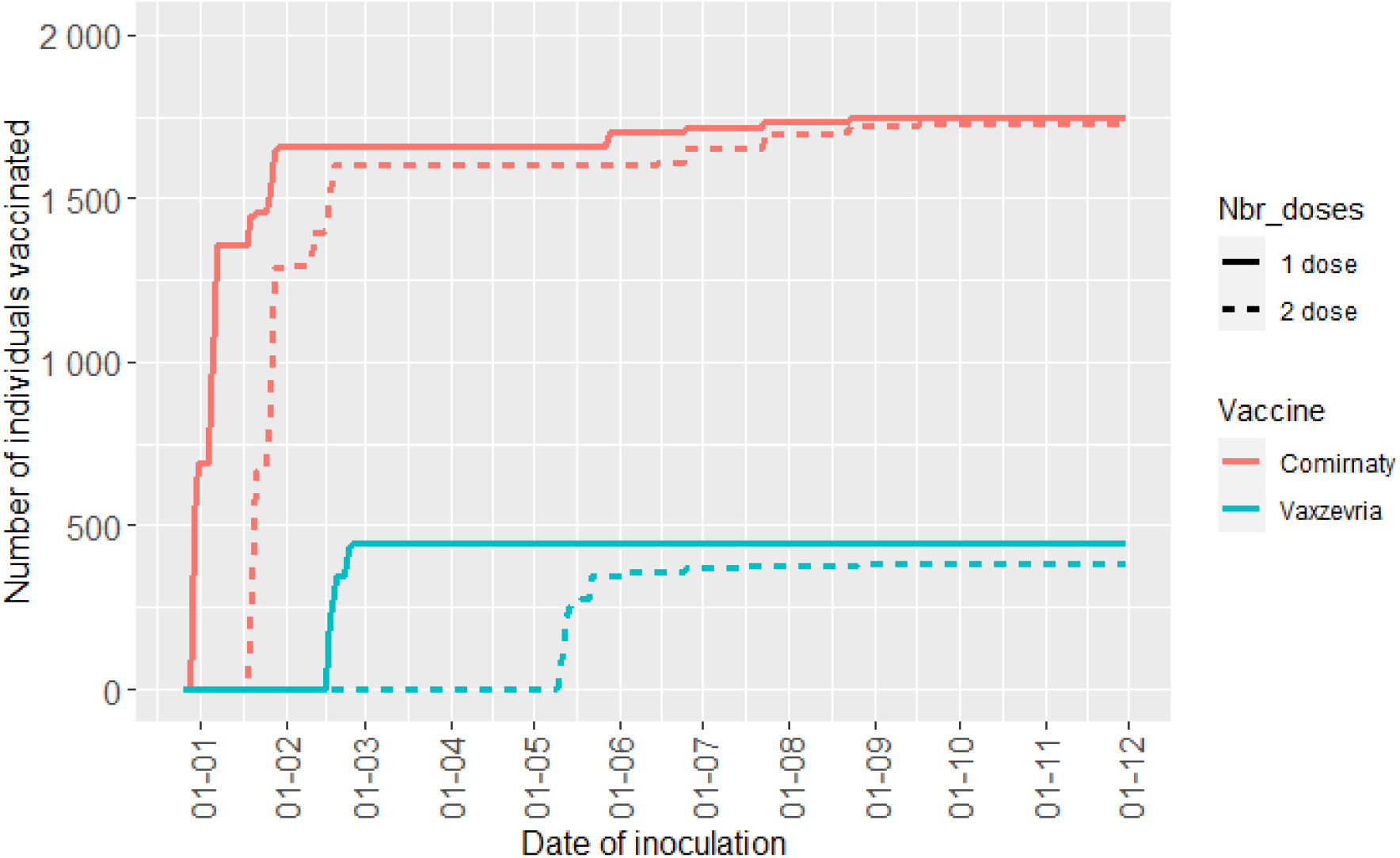
Evolution of vaccination status of the participants over the study period (Notes: Participants who reported to take the Spikevax vaccine (n=2) are not represented in this figure. For the situations of heterologous vaccination, the first dose brand was considered to define the vaccine brand).

Most incident cases of SARS-COV-2 infection (positive RT-PCR symptomatic cases) occurred at the beginning of the study (January 2021) (**Figure 2**).

**Figure 2.**
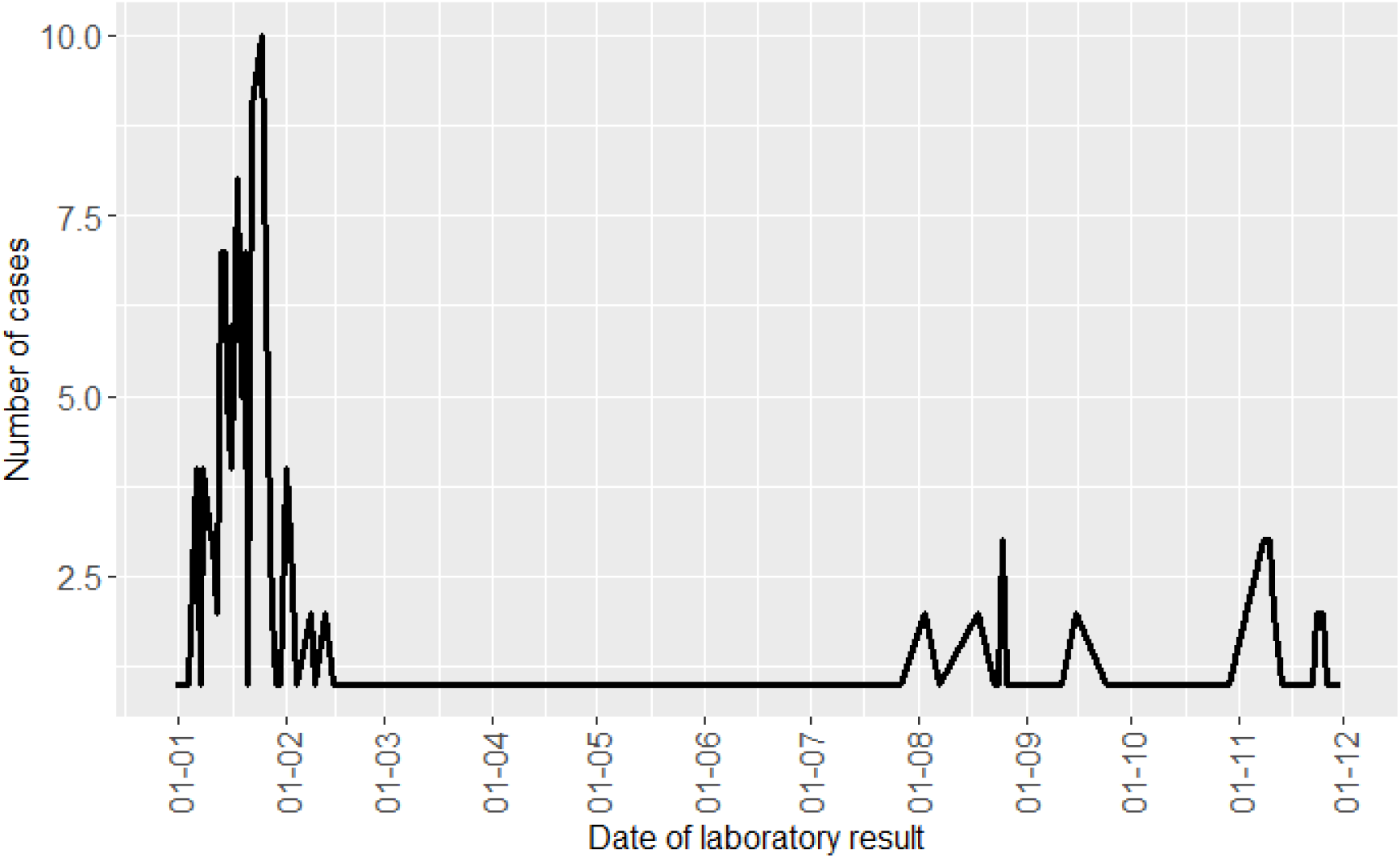
Evolution of the events (positive RT-PCR symptomatic cases) during the study period.

### Vaccine effectiveness against symptomatic COVID-19

Vaccine effectiveness against symptomatic infection was 78.0% (95%CI: 56.8 to 88.8) for partially vaccinated HCW and 73.9% (95%CI: 26.2 to 90.8) for fully vaccinated HCW (**Table 2**). The difference between these two estimates were not statistically significant.

**Table 2.** Vaccine effectiveness against symptomatic COVID-19.

| Vaccine Status | Person-years | Events (n) | Rate* | Rate Ratio (95% CI) | Confounder-adjusted HR (95% CI) | VE (95%CI) |
| --- | --- | --- | --- | --- | --- | --- |
| Unvaccinated | 238 | 83 | 348.7 | - | - | - |
| Partially | 224 | 24 | 107.1 | 0.36 (0.22 to 0.59) | 0.22 (0.11 to 0.43) | 78.0 (56.8 to 88.8) |
| Fully Vaccinated | 1488 | 64 | 43.0 | 0.15 (0.10 to 0.20) | 0.26 (0.09 to 0.74) | 73.9 (26.2 to 90.8) |
| **Total** | **1950** | **171** | **-** | **-** | **-** | **-** |

\*COVID-19-infection rates per 1,000 person-years. Abbreviations: HR: hazard ratio; VE: vaccine effectiveness.

## Discussion

Our study suggests high levels of protection against symptomatic SARS-CoV-2 infection in Portuguese HCW, conferred by full vaccination schemes of Comirnaty, and Vaxzevria. To our knowledge, this is one of the first studies to estimate VE in HCW, including during the Delta variant phase. A similar cohort study, still in pre-print, also performed in HCW from the United States of America found a similar estimate of VE (82.3%, 95% CI: 75.1–87.4%) considering the period between December 2020 and September 2021 (20). Other studies of HCW have already been published but they are mostly referred to a period of time not comparable to the present study (previous to Delta variant predominance) and estimates tend to be higher (above 80%) than the estimate obtained in the present study (12–14).

One of the limitations of our study is related to the sample size and small number of unvaccinated individuals. In fact, the relatively small sample size of the HCW cohort (n=2213), the high vaccine coverage and the few events observed during the study period contributed to the low precision of the estimates. Moreover, most of the events occurred in a restricted period of time while no events were observed in most of the follow-up weeks. However, the distribution of events over time was comparable to the epidemic curve observed in the Portuguese community aged between 18-70 years old for the same period (21). On the other hand, outcome measurement was conducted using the same informatics platform for all participant HCW, and so differential measurement of outcome is not plausible.

In addition, this study did not include variables related with adherence to vaccination of other preventive behaviours and this may have resulted in residual confounding, as the three categories of vaccination status may be associated with those behaviours or according to other individual HCW characteristics that may be associated with either vaccination uptake or risk of infection.

Larger cohort studies of HCW with long follow-up periods relied on weekly symptoms reporting, which result in a high burden for the participants and are difficult to maintain. One possible solution to obtain more robust vaccine effectiveness estimates in this risk group is to use cohorts based on electronic registries as previously performed for other risk groups in Portugal (19). The present work did not use electronic registries because electronic registries also poses some limitations, namely in obtaining important adjustment data in hospital context, such as occupational exposures. Moreover, it would imply the full identification of HCW within the databases, something that is not yet operationalized.

Finally, due to the small sample size related limitations, it was not possible to estimate VE according to brand of vaccine or according to other individual HCW characteristics. This cohort study will continue to monitor VE in HCW during the winter 2021/2022 and efforts will be made to include HCW from other Portuguese hospitals in order to increase the precision of VE estimates in this risk group to SARS-CoV-2 infection.

## Conclusion

This cohort study suggests a high VE for COVID-19 vaccine effectiveness against symptomatic SARS-CoV-2 infection in Portuguese HCW, which is in concordance with previous studies from other countries. This cohort study will continue to monitor VE in HCW during the winter 2021/2022.

## Data Availability

All data produced in the present study are available upon reasonable request to the authors

## Funding

This study was funded by ECDC (Contract ECD.11486 “Developing an infrastructure and performing vaccine effectiveness studies for COVID-19 vaccine in the EU/EEA” Lot3 (HCW)).

## Acknowledgments

Authors are grateful to all healthcare workers from hospitals who accepted to participate in this study. Author are also grateful to members of the research group of the cohort namely: Alexandra Lima Roque (Centro Hospitalar de Lisboa Ocidental), Ana Catarina Dias (Centro Hospitalar Tondela-Viseu), Ana Paula Rodrigues (Instituto Nacional de Saúde Doutor Ricardo Jorge), André Peralta Santos (Direção-Geral da Saúde), Artur Paiva (Centro Hospitalar Universitário de Coimbra), Carlos Penha Gonçalves (Instituto Gulbenkian de Ciência), Elvira Perea (Centro Hospitalar de Lisboa Ocidental), Filipe Pimenta (Centro Hospitalar Tondela-Viseu), Inês Costa (Instituto Nacional de Saúde Doutor Ricardo Jorge), Isabel Antunes (Centro Hospitalar Universitário de Coimbra), João Gonçalves (Faculdade de Farmácia da Universidade de Lisboa), João Paulo Gomes (Instituto Nacional de Saúde Doutor Ricardo Jorge), Jocelyne Demengeot (Instituto Gulbenkian de Ciência), Lígia Antunes Gonçalves (Instituto Gulbenkian de Ciência), Lucília Araújo (Centro Hospitalar Universitário de Coimbra), Nuno Verdasca (Instituto Nacional de Saúde Doutor Ricardo Jorge), Patrícia Conde (Instituto Nacional de Saúde Doutor Ricardo Jorge), Raquel Guiomar (Instituto Nacional de Saúde Doutor Ricardo Jorge), Rui Pedro Lopes (Centro Hospitalar Tondela-Viseu), Sandra Martins (Direção-Geral da Saúde), Vânia Pacheco (Centro Hospitalar Universitário de Coimbra).

## Notes

### Competing Interest Statement

The authors have declared no competing interest.

### Author Declarations

All data were anonymised before statistical analysis by the occupational health services of each hospital. This study protocol was approved by the ethics committee of the Instituto Nacional de Saude Doutor Ricardo Jorge (date of approval. 16/04/2021), and by the ethics committees of the two participating hospitals, namely by the ethics committee of CHTV (Ref.06/16/04/2021, date of approval 16/04/2021) and CHLO (Ref. 2141, date of approval 19/05/2021).

